# Utility of 18F-Flurpiridaz PET Relative Flow Reserve in Differentiating Obstructive from Nonobstructive Coronary Artery Disease

**DOI:** 10.1101/2025.06.11.25329454

**Authors:** Diana M. Lopez, Dan Huck, Sanjay Divakaran, Jenifer M. Brown, Brittany Weber, Mark Lemley, Valerie Builoff, Aakash Shanbhag, Zhou Lan, Christopher Buckley, Mouaz H. Al-Mallah, Sharmila Dorbala, Ron Blankstein, Piotr Slomka, Marcelo F. Di Carli

**Affiliations:** Cardiovascular Imaging Program, Departments of Medicine and Radiology, Brigham and Women’s Hospital, Harvard Medical School, Boston, MA, USA; Department of Medicine Cedars-Sinai, Los Angeles, California, USA; Department of Radiology, Brigham and Women’s Hospital, Harvard Medical School, Boston, MA, USA; Center for Clinical Investigation, Channing Division of Network Medicine, Brigham and Women’s Hospital, Harvard Medical School, Boston, MA, USA; GE Healthcare, Pharmaceutical Diagnostics, Buckinghamshire, UK; Houston Methodist DeBakey Heart and Vascular Center, Houston, Texas, USA

## Abstract

**Background:** Absolute quantification of myocardial blood flow (MBF) on PET perfusion imaging improves the identification of coronary artery disease (CAD). However, distinguishing MBF impairment due to obstructive CAD from nonobstructive CAD remains challenging. We aimed to evaluate the incremental diagnostic value of PET-derived relative flow reserve (RFR) in the diagnosis of obstructive CAD.

**Methods:** This is a post hoc analysis of the multicenter phase-III trial of ^18^F-flurpiridaz PET (NCT01347710). Patients with available MBF quantification were included. Reduced stress MBF (sMBF) was defined as sMBF below the median (2.2 mL/min/g). Obstructive CAD on quantitative invasive coronary angiography (ICA) was defined as ≥70% stenosis. RFR was calculated as a ratio of the minimal segment sMBF over the highest reference vascular territory sMBF. RFR performance for predicting obstructive CAD was evaluated through Receiver Operating Characteristic (ROC) analysis and the net reclassification index (NRI) of multivariable regression models.

**Results:** The study included 231 patients (71% male; 56% with established CAD) drawn from the original cohort of 755 trial participants. No patients had three-vessel CAD. In a per vessel-based analysis, 82% of vessels with reduced sMBF had no obstructive CAD on ICA. RFR was significantly lower for vessels with obstructive CAD (0.55 vs 0.80, p<0.0001). In vessels with reduced sMBF, RFR was independently associated with obstructive CAD even after accounting for sTPD and MFR (OR 3.08, 95% CI: 1.49–6.38; p = 0.002). While the addition of RFR did not significantly improve discrimination (AUC 0.806 vs. 0.822, p = 0.11), it significantly improved reclassification of vessels with and without obstructive CAD (NRI: 0.93, p < 0.0001).

**Conclusions:** RFR provides complementary diagnostic information beyond existing PET parameters and may help refine the diagnosis of obstructive CAD in patients with reduced flows.

**Clinical Perspective:** A major diagnostic dilemma in cardiac PET/CT perfusion imaging is determining whether reductions in stress myocardial blood flow and/or myocardial flow reserve are caused by obstructive or nonobstructive coronary artery disease (CAD), leading to uncertainty about whether invasive angiography is needed. Our study demonstrates that incorporating PET-derived relative flow reserve (RFR) adds meaningful diagnostic information beyond existing PET perfusion and flow parameters. RFR does not substantially increase overall discrimination between obstructive and nonobstructive CAD, but it significantly improves reclassification of individual cases, indicating that RFR can help refine decision-making, particularly in borderline cases. These data suggest that selective integration of RFR with existing PET metrics could improve patient selection for invasive procedures and guide more targeted medical therapy for nonobstructive CAD. Future research is needed to confirm these findings across a broader patient population, including higher-risk cohorts and women, as well as with other PET radiotracers.

## Introduction

A major diagnostic dilemma in cardiac PET/CT perfusion imaging is determining whether reductions in stress myocardial blood flow (sMBF) and/or myocardial flow reserve (MFR) are caused by obstructive or nonobstructive coronary artery disease (CAD) – a distinction that has significant implications for additional testing, including referral to invasive coronary angiography (ICA). Cardiac PET/CT perfusion imaging integrates multiple physiologic imaging parameters, which improves the identification and risk stratification of coronary artery disease (CAD).^1, 2^ However, reductions in sMBF and MFR can occur in the absence of focal obstructive epicardial disease.^2–4^ This is attributed to the presence of diffuse nonobstructive epicardial atherosclerosis and coronary microvascular disease, which are increasingly prevalent with the rise of cardiometabolic disease.^5–8^ From a clinical perspective, the presence of regional or global reductions in sMBF and/or MFR in patients with normal or mildly abnormal myocardial perfusion creates uncertainty regarding the underlying cause of the flow abnormalities. Consequently, additional tools are needed to differentiate classic obstructive CAD from nonobstructive atherosclerosis.

Prior studies have investigated the diagnostic utility of the PET-derived relative flow reserve (RFR), a non-invasive measure that compares sMBF in a myocardial region of interest to a normal reference region, thereby calculating a flow ratio to understand whether the stenosis is hemodynamically significant.^9–13^ Coronary flow and pressure are linearly related under maximal hyperemic conditions; thus, the RFR could function as a non-invasive surrogate of the fractional flow reserve measured using a pressure wire in the catheterization laboratory. Prior studies have shown that RFR, when used as a standalone metric, was not superior to sMBF or MFR for detecting flow-limiting CAD.^9–11^ However, the complementary role of RFR in distinguishing PET myocardial flow abnormalities caused by angiographically obstructive CAD from those associated with diffuse nonobstructive atherosclerosis remains unclear. Therefore, our objective was to investigate the incremental value of RFR when integrated with existing quantitative PET parameters in symptomatic patients who underwent Flurpiridaz PET and invasive angiography as part of a blinded prospective phase III study.^14^

## Methods

The data that support the findings of this study are available from the corresponding author upon reasonable request.

### Study Population

Patients were drawn from the first Flurpiridaz myocardial perfusion PET phase III trial (NCT01347710).^14^ Briefly, participants were ≥18 years old and had to be referred for clinically indicated ICA. Major exclusion criteria included acute coronary syndrome or percutaneous coronary intervention within 6 months, stroke within 3 months, prior coronary artery bypass graft surgery, symptomatic valvular disease, significant congenital heart defects, New York Heart Association functional class III-IV heart failure, nonischemic cardiomyopathy, and history of heart transplantation. Pre-test probability of CAD was assessed per the ACC/AHA guidelines for exercise testing.^15^ PET studies were performed within 60 days of ICA. Dynamic imaging for absolute flow quantification was optional and performed at the discretion of the recruitment sites. Institutional Review Board approval was obtained at each study site and written informed consent was obtained from all the trial patients.

### Invasive Coronary Angiography

ICA was performed in accordance with each site’s clinical protocol. Images were evaluated by the designated angiography core laboratory (Boston Clinical Research Institute, Boston, MA, USA) for blinded quantitative percent diameter coronary stenosis measurements (QCAPlus, Sanders Data Systems, Palo Alto, CA USA). For this post hoc study, obstructive CAD was defined as ≥ 70% stenosis in the left anterior descending artery (LAD), left circumflex artery (LCX), right coronary artery (RCA), or their major branches, and ≥ 50% stenosis in the left main artery (LM). Obstructive CAD in the LM was categorized as 2-vessel CAD.

### Flurpiridaz Myocardial Perfusion PET/CT Imaging Protocol

PET myocardial perfusion imaging (MPI) was performed as previously described.^14, 16^ Briefly, vasodilator stress was performed with regadenoson, adenosine, or dipyridamole in accordance with each study site’s protocol. Dynamic PET MPI acquisition at rest and at peak stress was performed following intravenous bolus injections of ^18^F-flurpiridaz at doses of 2.7 ± 0.2 mCi and 5.9 ± 0.3 mCi, respectively, with a minimum of a 30-minute interval between rest and stress injections.

#### Motion Correction

Motion was corrected manually for each frame at stress and rest to align the myocardial tracer uptake in each frame with myocardial contours, as previously reported.^17, 18^ For each frame in each dataset, the operators shifted the image in relation to the static LV myocardial contours from the static image in the three principal axes (x: lateral to septal, y: anterior to inferior, and z: apex to base). The magnitude of motion was then assessed across all patients in the direction of each of the axes. The frequency of motion shift ≥ 5 mm was calculated.

#### Residual Activity Subtraction

Residual activity correction (RAC) was applied to the stress dynamic images to account for the bias introduced by the residual 18F-flurpiridaz activity from the rest injection, as previously described.^17, 19^

### Relative Perfusion Image Analysis

For semiquantitative perfusion analysis, static images were created by summing the dynamic PET images after the initial 2 minutes. Perfusion images were processed in batch mode with dedicated software (QPET, Cedars-Sinai Medical Center, Los Angeles, CA).^20^ Normal database comprised patients with normal studies from the phase II 18F-flurpiridaz trial.^21^ The total perfusion deficit (TPD) at rest and stress was computed as previously described for the entire left ventricle and each coronary vascular territory (LAD, LCX, RCA).^22^ Ischemic TPD was also calculated and defined as the difference between stress and rest TPD. Abnormal stress total perfusion deficit (sTPD) was defined as ≥7%, a threshold previously found to provide the optimal diagnostic operating point.^23^

### Quantification of Myocardial Blood Flow and Flow Reserve

Myocardial blood flow and flow reserve maps were generated from the dynamic image series with the QPET software using a 2-compartment kinetic model.^19^ Quantitative MBF in each coronary territory was estimated using the tracer uptake kinetics within the first 90 seconds post-tracer injection. The spillover fraction from the blood pool to the myocardium plus the vascular volume of distribution was approximated as 1.0 minus the recovery coefficient of the corresponding myocardial sample. MBF was obtained by assuming a first-pass extraction fraction of 0.94.^24^ Myocardial flow reserve polar maps were computed by dividing the stress and rest MBF in each polar map sample. Average MBF and MFR values were quantified for the entire left ventricle, each coronary vascular territory (LAD, LCX, RCA), and for each of the 17 myocardial segments in accordance with AHA standardized segmentation model.^25^ MBF measurements were not adjusted for rate pressure product. Since the threshold for an abnormal ^18^F-flurpiridaz sMBF has not been established in larger studies, we defined reduced per vessel sMBF as values falling below the group median. Severely reduced sMBF was defined as < 1.5 ml/min/g.^26^ Impaired MFR was defined as MFR <2.0.^27, 28^

### Relative Flow Reserve

The relative flow reserve was calculated for each vessel by dividing the lowest segmental sMBF^12^ in each vessel territory by the average sMBF in the territory with the highest flow value, which was considered the reference territory (Supplementary Figure 1). This approach is consistent with previously published RFR methodology and allows for patient-specific normalization based on the most preserved vascular territory.^9, 10, 12^ The apical 17th segment was excluded in the estimation of LAD RFR. The process was automated to determine the lowest segmental and reference vascular flows without user selection and the reviewer assessing these values was blinded to the presence or absence of obstructive CAD on ICA.

### Statistical Analysis

Categorical variables are reported as frequencies with percentage and compared using chi-squared test. Continuous variables are presented as mean ± standard deviation or median with interquartile range (IQR), and compared using Wilcoxon rank-sum and Kruskal-Wallis tests, as appropriate.

Per-vessel analyses were performed by clustering at the patient level to account for within-patient correlation. Each patient contributed up to three matched PET and ICA measurements (LAD, LCX, and RCA). Univariate receiver operating characteristic (ROC) analysis was performed for each PET parameter and differences in area under the curve (AUC) were compared using DeLong’s test, with 95% confidence intervals (CI) estimated via clustered bootstrapping. Predictive margins were plotted from a generalized estimating equations (GEE) model with patient-level clustering. The optimal RFR threshold was selected with Youden’s index. To evaluate the incremental diagnostic value of RFR, multivariable logistic regression models with GEE to account for clustering. Models included adjustment for age, sex, and BMI. Interaction terms between the PET parameters were retained only if statistically significant. ROC and continuous net reclassification index (NRI) further assessed model performance, with CI and comparisons derived from clustered bootstrapping. All tests were 2-sided, and p <0.05 was considered statistically significant. Analyses were performed using Stata/BE 17.0 (Statacorp) and R version 4.5.0 (R Foundation for Statistical Computing).

## Results

### Study Population

Absolute flow quantification was available for 275 of the 755 patients in the trial. Of these, 231 patients were included in this post hoc study after excluding studies with limited quality flow data. Reasons for quality control failure included missing or corrupted dynamic images (n = 5), unavailable heart rate and blood pressure at rest (n = 1), or abnormal left ventricular (LV) input curves (n = 38), defined as flat curves, absent peaks, or multiple peaks. Patient characteristics are summarized in **Table 1**. The mean age was 61.9 years (± 9.4), and the majority of the study participants were white (78%), male (71%), had established CAD (56%) and a high burden of cardiometabolic risk factors. Most patients were symptomatic (88%), with typical chest pain being the most common reason for ICA referral (45%).

**Table 1.**
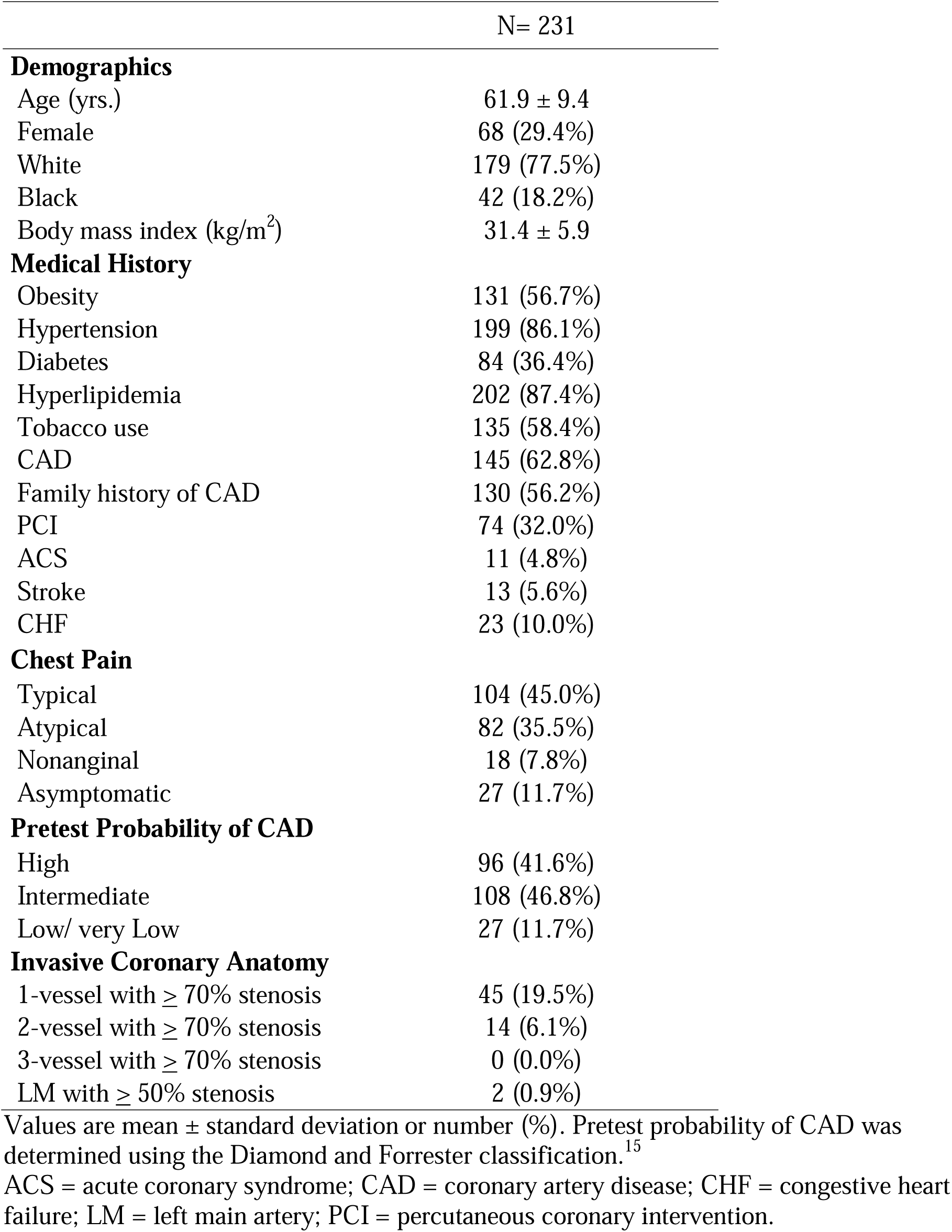
Patient characteristics.

### PET and ICA Results

Per-patient ICA and PET results are summarized in **Table 1** and **Supplementary Table 1**; per-vessel results are summarized in **Table 2**. Despite a high burden of symptoms and cardiometabolic risk factors, only 25.5% of patients (59 out of 231) had obstructive CAD. Multi-vessel obstructive CAD was uncommon: less than 10% had 2-vessel disease and none had 3-vessel obstructive CAD. The median sMBF and MFR were 2.2 mL/min/g (IQR 1.7–2.8) and 2.9 (IQR 2.2–3.7), respectively. These values were consistent at the global and per-vessel levels. Angiographically obstructive CAD was present in 10.5% (73/693) of vessels. RFR was lower in vessels with obstructive CAD when compared to nonobstructive vessels (0.55 vs 0.80, p <0.0001).

**Table 2.** Per-vessel PET and ICA Results.

|  | ALL Vessels<br>(n = 693) | Nonobstructive CAD<br>(n = 620) | Obstructive CAD<br>(n = 73) | p-value |
| --- | --- | --- | --- | --- |
| **sMBF (ml/min/g)** | 2.22 (1.68, 2.79) | 2.32 (1.82, 2.88) | 1.38 (1.07, 1.77) | <0.0001 |
| **MFR** | 2.93 (2.23, 3.74) | 3.00 (2.35, 3.81) | 1.86 (1.56, 2.53) | <0.0001 |
| **Stress TPD ≥ 7%** | 129 (18.6%) | 83 (13.4%) | 46 (63.0%) | <0.0001 |
| **RFR** | 0.79 (0.66, 0.90) | 0.80 (0.70, 1.00) | 0.55 (0.43, 0.70) | <0.0001 |

Per-vessel values from 231 patients. Values are median (IQR) or number (%). P-values were derived from clustered bootstrapping. Obstructive CAD was defined as stenosis ≥ 70% (or LM ≥ 50%). ICA = invasive coronary angiogram; MBF = myocardial blood flow; MFR = myocardial flow reserve; PET = positron emission tomography; RFR = relative flow reserve; TPD = total perfusion deficit.

### Obstructive CAD by PET Flow and Flow Reserve

To investigate the relationship between per vessel sMBF and angiographic stenosis in this study population, we examined the distribution of obstructive CAD by sMBF and MFR (**Figure 1**). The majority of vessels with angiographically obstructive CAD had reduced sMBF (88%, p <0.001). Of these, the largest proportion (53%) had concordantly reduced sMBF and MFR (p <0.001). However, most vessels with reduced sMBF (82%, 282/346), including those with concordantly reduced MFR (61%, 60/99), had no obstructive CAD on ICA.

**Figure 1.**
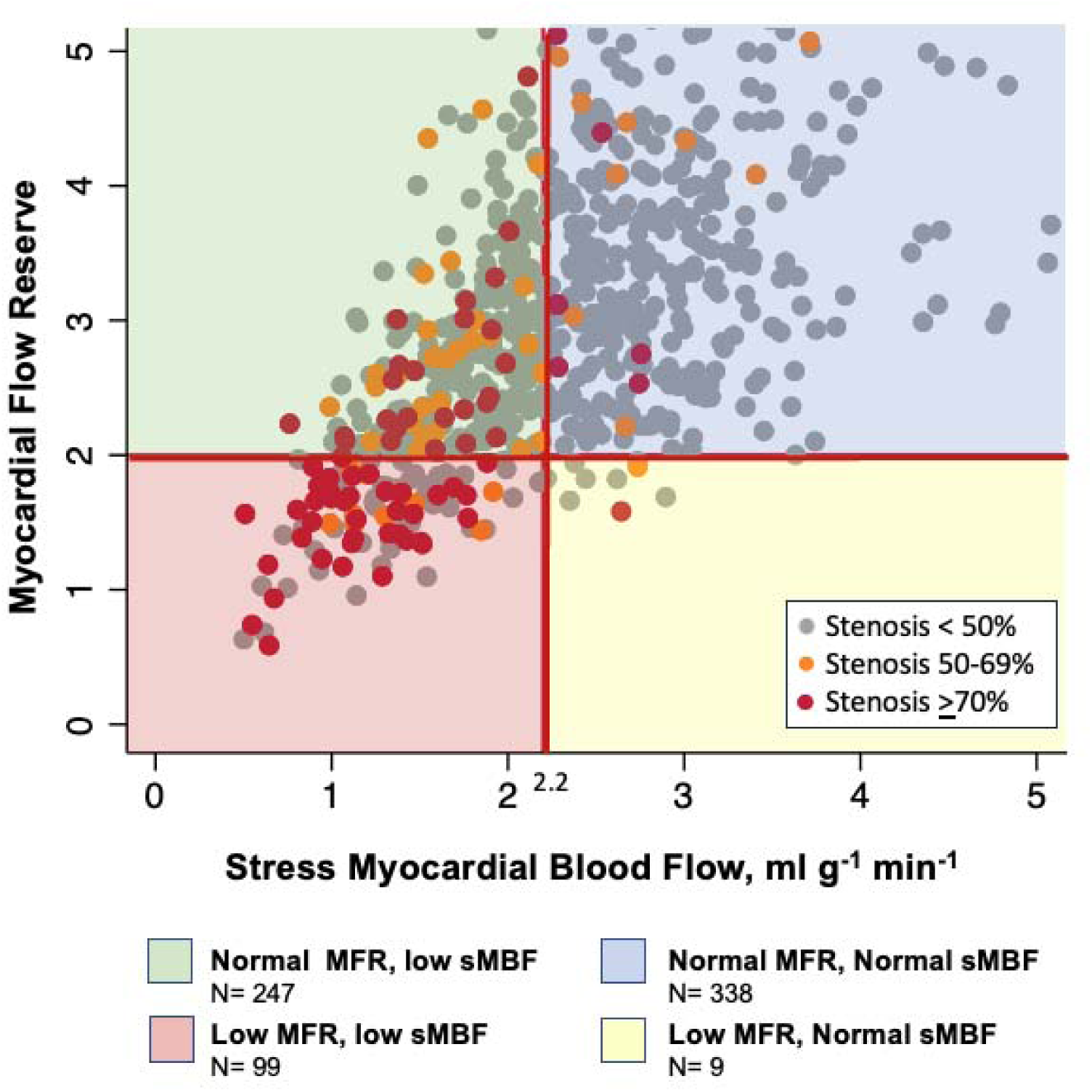
Per-vessel distribution of CAD by PET flow groups. Scatter plot of myocardial flow reserve and stress myocardial blood flow for each PET vessel territory of 693 vessels (231 patients). Dot color indicates severity of epicardial stenosis by invasive angiography in the corresponding vessel.

### Integration of Flow and Perfusion Metrics

Given that the clinical interpretation of stress PET MPI relies on both flow and perfusion metrics, we examined the distribution of obstructive CAD when stratifying vessels by both abnormal sMBF and abnormal sTPD (>7%) (**Figure 2**). In vessels with normal sMBF, an abnormal sTPD did not significantly differentiate vessels with and without obstructive CAD (p = 0.298), reflecting the low prevalence of obstructive disease (n=9 vessels) in this subgroup. In vessels with reduced sMBF, an abnormal sTPD was associated with a higher proportion of vessels with obstructive CAD compared to a normal sTPD (43% vs 8%, respectively, p <0.0001); however, notably more than half of the vessels with abnormal sTPD and reduced sMBF had nonobstructive CAD on ICA.

**Figure 2.**
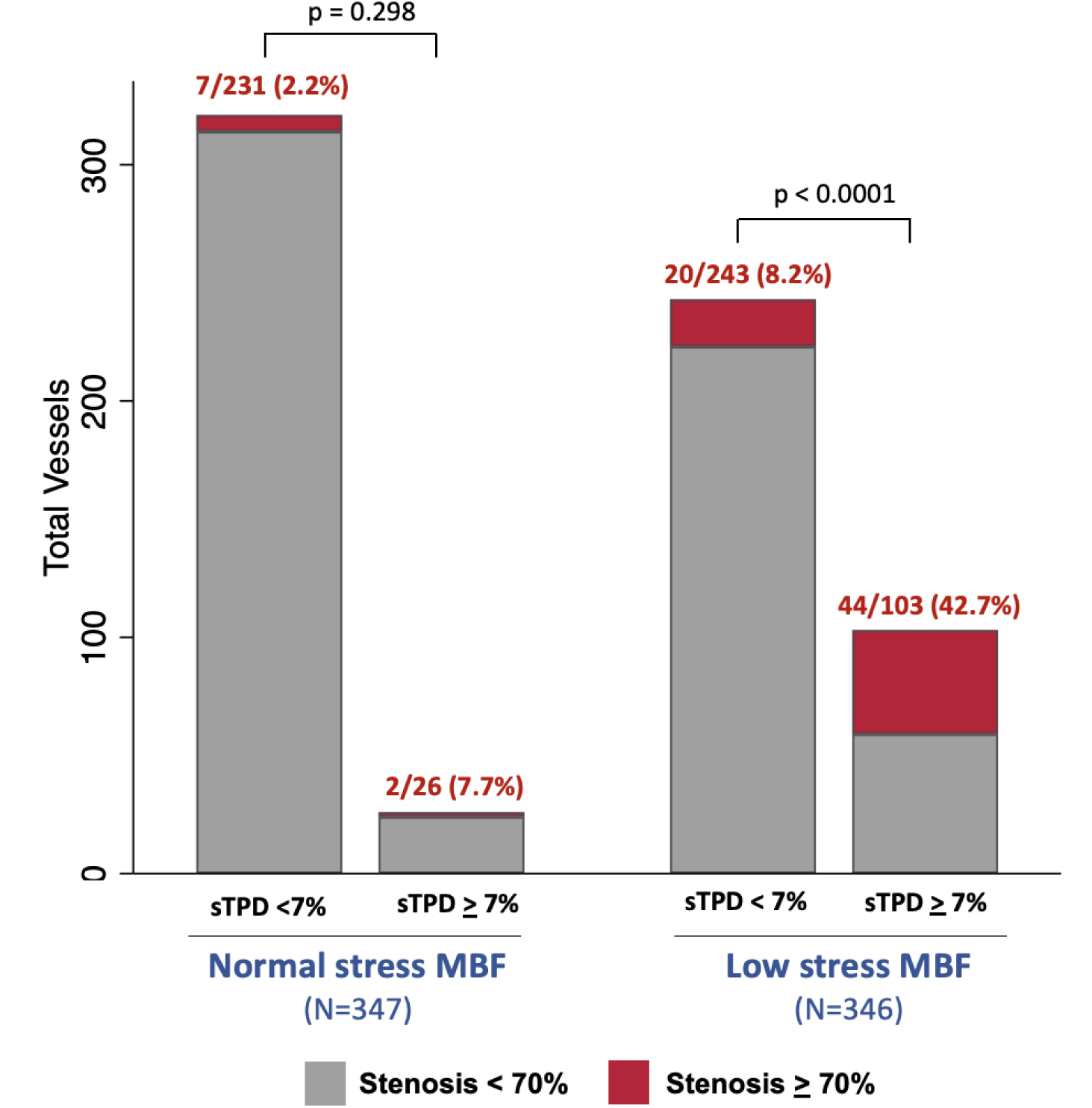
Per-vessel obstructive CAD by abnormal stress MBF and stress TPD. Data from 693 vessels (231 patients). Bar heights reflect group size; percentages represent the proportion of vessels with obstructive CAD (≥70% stenosis or ≥50% LM) in each group. P-values derived from chi-square tests with clustered bootstrapping. MBF = myocardial blood flow; TPD = total perfusion deficit.

### RFR and Stenosis Severity

We examined the distribution of RFR across stenosis severities (**Figure 3A**). The median RFR for all vessels was 0.79 (IQR 0.66–0.90). RFR declined with increasing stenosis severity, dropping significantly to 0.55 (IQR 0.43–0.70) with severely stenotic lesions (p = 0.001). RFR differences between obstructive vs nonobstructive vessels were evident across all three vascular territories (**Figure 3B**). Standalone per-vessel discriminatory performance of RFR and the other PET metrics is shown in **Supplementary** Figure 2. The optimal RFR cutoff for the diagnosis of obstructive CAD was 0.64 with sensitivity and specific of 67% and 84% respectively. There was a nonsignificant trend of higher obstructive CAD probability with lower RFR at a given MBF or sTPD **(Supplementary** Figure 3).

**Figure 3.**
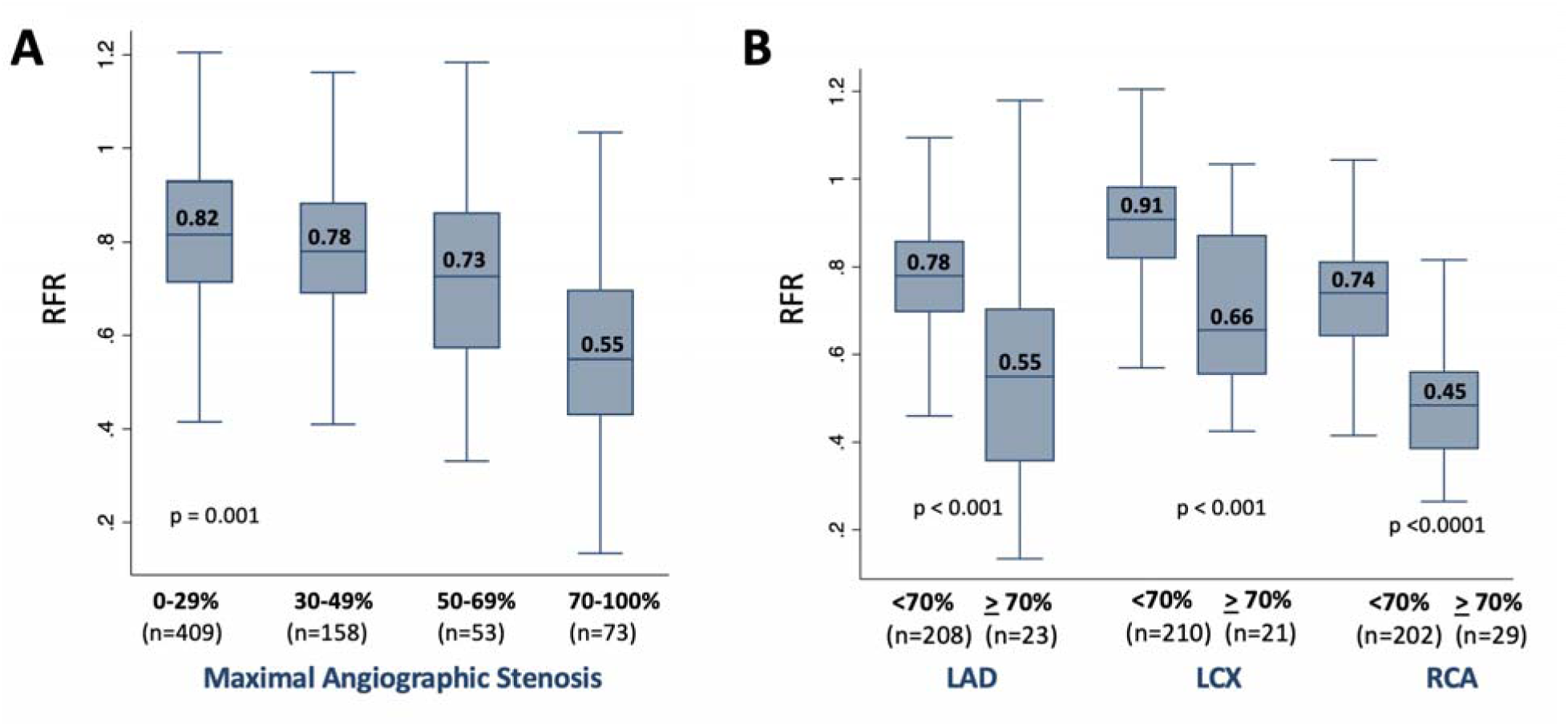
RFR with angiographic stenosis. All data are from 693 vessels in 231 patients. **(A)** RFR by increasing severity of angiographic stenosis. P-value from clustered bootstrapping. **(B)** RFR distribution in obstructive versus nonobstructive vessels stratified by vascular territory. Boxes = IQR (25th–75th percentiles) with median values shown; whiskers extend to the minimum and maximum values (excluding outliers). LAD = left anterior artery; LCX = left circumflex artery; RCA = right coronary artery; RFR = relative flow reserve.

### Incremental Diagnostic Value of RFR

To investigate the incremental value of RFR, multivariable logistic analysis was performed on vessels with reduced sMBF (**Table 3)**. Even after adjusting for clinical covariates, sTPD, and MFR, RFR was independently associated with a threefold higher odds of obstructive CAD (OR 3.08, 95% CI 1.49–6.38, p=0.002). The addition of RFR improved discrimination between obstructive and nonobstructive CAD when compared to using either sTPD alone (AUC 0.763 vs 0.794, p = 0.010; **Figure 4**) or to MFR alone (AUC 0.718 vs 0.801, p < 0.002; **Supplementary** Figure 4). However, RFR did not significantly improve discrimination when added to both sTPD and MFR (AUC 0.806 vs 0.822, p = 0.11; **Figure 4**). From a reclassification standpoint, however, the addition of RFR yielded a significant improvement over both sTPD alone (NRI of 0.958, 95% CI: 0.710–1.206, p < 0.0001) and a combination of both sTPD and MFR (NRI of 0.927, 95% CI: 0.665–1.188, p < 0.0001) (**Table 4**).

**Figure 4.**
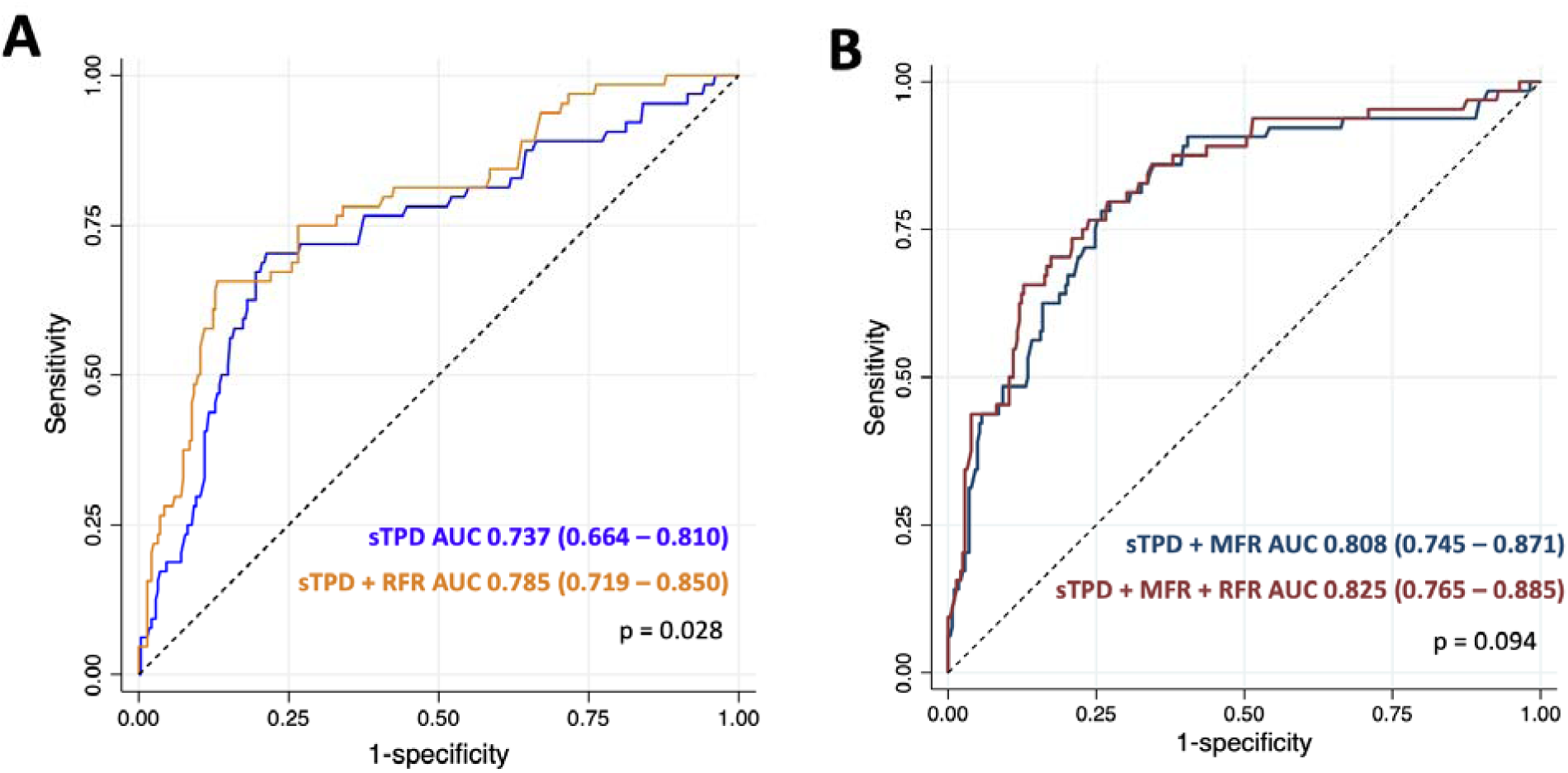
Per-vessel Incremental Discriminatory Value of RFR for Identifying Obstructive CAD in Vessels with Reduced Stress MBF. ROC curves comparing (A) sTPD alone versus sTPD and RFR, and (B) MFR alone versus MFR and RFR. Models adjusted for age, BMI, and sex. P-values and 95% confidence intervals derived from clustered bootstrapping in an analysis of 346 vessels from 143 patients. MBF = myocardial blood flow; MFR = myocardial flow reserve; RFR = relative flow reserve; ROC = receiver operating characteristic; sTPD = stress total perfusion deficit.

**Table 3.**
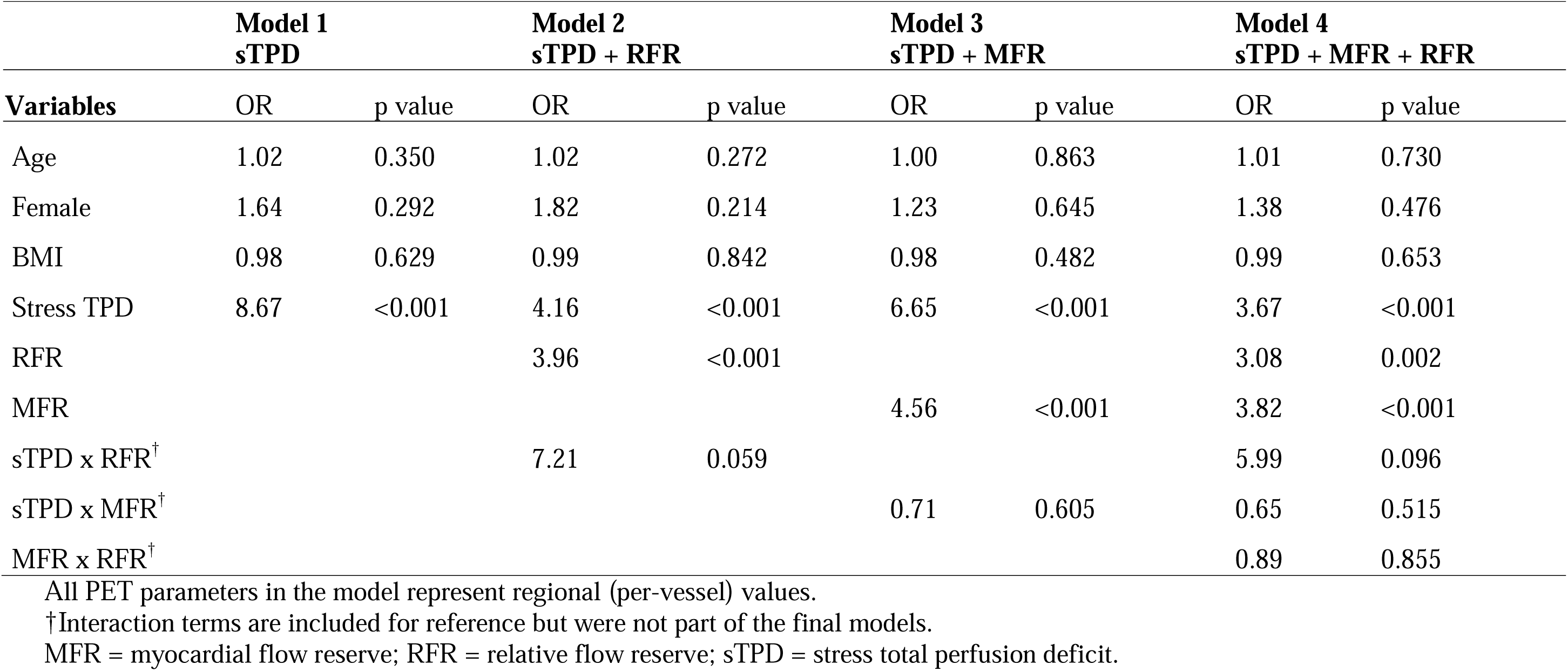
Multivariable regression models for detection of obstructive CAD in vessels with reduced stress MBF.

| **Variables** | **Model 1**<br>**sTPD** |  | **Model 2**<br>**sTPD + RFR** |  | **Model 3**<br>**sTPD + MFR** |  | **Model 4**<br>**sTPD + MFR + RFR** |  |
| --- | --- | --- | --- | --- | --- | --- | --- | --- |
|  | OR | p value | OR | p value | OR | p value | OR | p value |
| Age | 1.02 | 0.350 | 1.02 | 0.272 | 1.00 | 0.863 | 1.01 | 0.730 |
| Female | 1.64 | 0.292 | 1.82 | 0.214 | 1.23 | 0.645 | 1.38 | 0.476 |
| BMI | 0.98 | 0.629 | 0.99 | 0.842 | 0.98 | 0.482 | 0.99 | 0.653 |
| Stress TPD | 8.67 | <0.001 | 4.16 | <0.001 | 6.65 | <0.001 | 3.67 | <0.001 |
| RFR |  |  | 3.96 | <0.001 |  |  | 3.08 | 0.002 |
| MFR |  |  |  |  | 4.56 | <0.001 | 3.82 | <0.001 |
| sTPD x RFR<sup>†</sup> |  |  | 7.21 | 0.059 |  |  | 5.99 | 0.096 |
| sTPD x MFR<sup>†</sup> |  |  |  |  | 0.71 | 0.605 | 0.65 | 0.515 |
| MFR x RFR<sup>†</sup> |  |  |  |  |  |  | 0.89 | 0.855 |

All PET parameters in the model represent regional (per-vessel) values.
<sup>†</sup>Interaction terms are included for reference but were not part of the final models.
MFR = myocardial flow reserve; RFR = relative flow reserve; sTPD = stress total perfusion deficit.

**Table 4.**
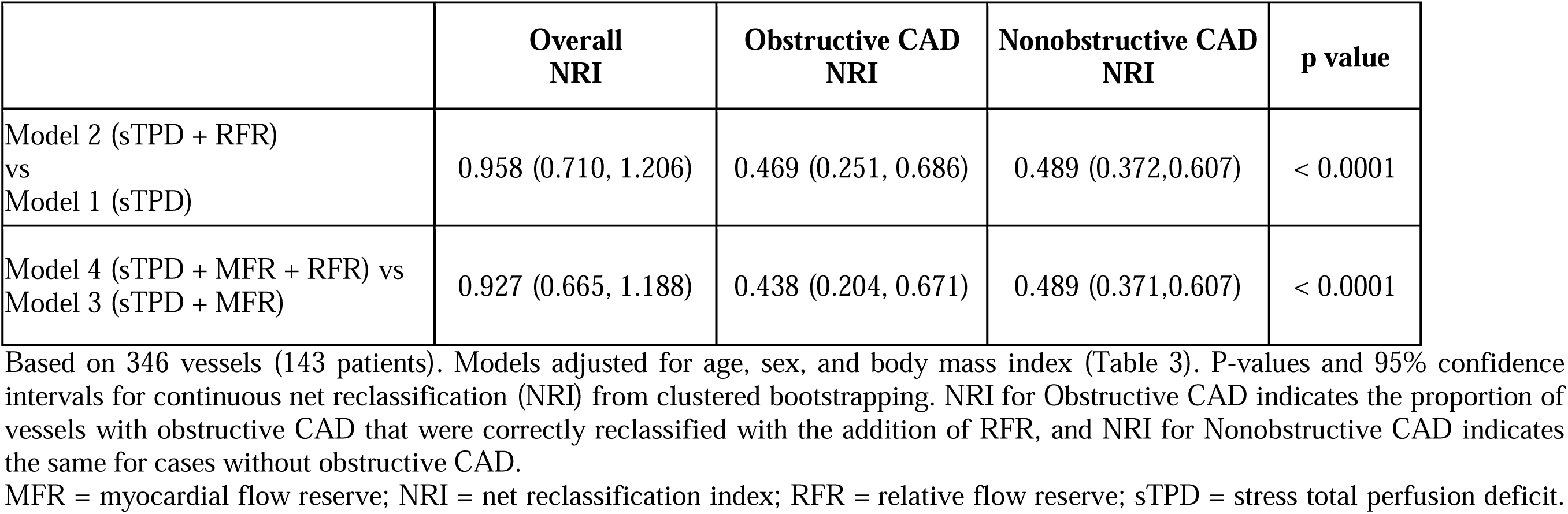
Reclassification of obstructive CAD in vessels with reduced stress MBF.

## Discussion

Accurate and reproducible MBF quantification is a key advantage of PET MPI over other cardiac imaging modalities. These flow measurements improve detection of flow-limiting CAD and risk stratification in all patient groups.^29–31^ Nevertheless, the interpretation of reduced flows often introduces diagnostic uncertainty as to whether they reflect underlying obstructive or nonobstructive CAD. The integrated assessment of sMBF and MFR was previously shown to identify differential risk for cardiac death.^32^ Our study builds on this by showing that concordantly reduced regional sMBF and MFR were associated with the highest prevalence of obstructive CAD. Yet, 82% of vascular territories with reduced sMBF—and 61% of those with concordantly impaired sMBF and MFR—had no obstructive CAD, highlighting the need for additional metrics to refine diagnostic certainty in patients with impaired PET flow parameters.

Lower flow impairment thresholds have been proposed to improve specificity for detecting obstructive CAD.^28^ Still, in our study, most vascular territories with severely reduced sMBF (<1.5 mL/min/g) did not have obstructive CAD on ICA. Abnormal perfusion (sTPD ≥7%) improved the differentiation between obstructive and nonobstructive CAD, but more than half of the vascular territories with abnormal sMBF and sTPD had nonobstructive CAD. These findings highlight the limitations of existing PET metrics. This is especially relevant with high extraction fraction tracers like ^18^F-flurpiridaz, ^13^N-ammonia, and ^15^O-water—as these agents are more sensitive flow limitations caused by moderate or diffuse disease—and emphasize the need for additional strategies.

In this context, our study offers new insights into the application of ^18^F-flurpiridaz PET-derived RFR for the diagnosis of obstructive CAD. We found that RFR was strongly associated with stenosis severity, with the lowest values in vessels with severe stenoses. Among vessels with reduced sMBF, RFR was independently predictive of obstructive CAD, even after accounting for perfusion and MFR (OR 3.08; p=0.002), suggesting that RFR provides distinct physiologic information. RFR did not significantly improve global discriminatory performance (AUC 0.806 vs 0.822, p = 0.11), but it improved vessel-level classification beyond sMBF and MFR for both obstructive and nonobstructive CAD cases. As such, RFR may be particularly useful for clarifying diagnostic uncertainty in patients with intermediate findings on sTPD or MFR.

While ROC-based test assessment is often the go-to approach, the AUC or c statistic can be relatively insensitive to incremental improvements in prediction, particularly when the baseline test already demonstrates good discrimination. Cook^33^ notes that even well-established predictors of CVD may have only a marginal impact on the c statistic individually, especially if the new metric is geared more toward refining individual risk prediction rather than overall discrimination. Kerr et al.^34^ similarly caution against relying solely on NRI-based assessment. Guided by these principles, this study did not rely on a single metric but rather evaluated the added value of RFR through multiple complementary approaches.

Past studies that evaluated the diagnostic utility of PET-derived RFR had mixed results.^9–13^ Stuijfzand et al.^9^ reported that RFR was not superior to sMBF or MFR in diagnosing obstructive CAD. Cho et al.^11^ showed that RFR declined more prominently with focal stenosis and remained relatively preserved in the setting of diffuse atherosclerosis compared to MFR, suggesting that RFR provides complementary physiologic insights. These findings highlight the importance of integrating multiple PET-derived parameters to comprehensively phenotype coronary physiology.

The integration of RFR requires an understanding how this metric aligns with both anatomical and hemodynamic assessments of stenosis severity. De Bruyne et al.^13^ demonstrated that RFR correlates better with invasive fractional flow reserve (FFR) than with percent diameter stenosis.

This may explain the wide RFR distribution in vessels with severe stenosis. Notably, this is not unique to RFR. Severe stenosis on ICA can be associated with substantial variability in PET-based sMBF and MFR,^26, 35, 36^ as well as invasive FFR.^37, 38^ Similarly, we observed that vessels without obstructive CAD often had RFR values well below 1.0—paralleling the phenomenon in which diffuse nonobstructive disease can also yield abnormal FFR values.^37–39^ This physiologic-anatomic discordance may have also impacted RFR’s ability to improve the ROC-based discrimination assessment, which essentially reflects how well a test separates two groups as distinct populations.

### Limitations

Our study has several limitations. Patients underwent routine clinically indicated ICAs and invasive FFR was not available. However, anatomical stenosis remains the most common metric for assessing lesion severity in clinical practice. MBF quantification was not performed in all trial participants and patients who received PCI at the time of ICA were excluded from the trial, potentially limiting generalizability. Raw ICA images were not available; therefore vessel assignment was based on standard territory definitions, which may not reflect individual patient anatomy. No patients had three-vessel obstructive CAD, so RFR’s utility in this group remains unknown. The cohort was mostly male, warranting further investigation of RFR performance in women. Finally, these findings apply to 18F-flurpiridaz PET-derived RFR; further studies are needed to assess RFR performance in other PET tracers like ^82^Rb-rubidium, as its lower extraction fraction may affect RFR performance and diagnostic thresholds.

### Conclusions

In patients with 1-to 2-vessel obstructive CAD, RFR is significantly reduced in vessels with severe stenoses and provides incremental diagnostic value in differentiating obstructive and nonobstructive CAD in vessels with reduced myocardial blood flow.

## Data Availability

The data underlying this study are proprietary and cannot be shared publicly. Access to the data may be granted by the clinical trial sponsor upon reasonable request.

## Acknowledgements

This work was conducted with support from Harvard Catalyst | The Harvard Clinical and Translational Science Center (National Center for Advancing Translational Sciences, National Institutes of Health Award UL 1TR002541).

## Supplementary Material

Figure S1

Figure S2

Figure S3

Figure S4

Supplementary Table 1

## Non-standard Abbreviations and Acronyms

CMD: coronary microvascular disease
sMBF: stress myocardial blood flow
MFR: myocardial flow reserve
RFR: relative flow reserve
sTPD: stress total perfusion deficit

