## Supplemental Figures and Table for "Utility of 18F-Flurpiridaz PET Relative Flow Reserve in Differentiating Obstructive from Nonobstructive Coronary Artery Disease"

<u>**Supplemental Material (For Publication)**</u>

### Supplemental Figures:

**Supplemental Figure 1. Relative Flow Reserve Method.** In this example, LAD RFR is calculated by dividing the lowest segmental stress MBF in the LAD territory (Segment 13) by average vascular stress MBF with the highest value (LCX). LAD = left anterior descending artery; LCX = left circumflex artery; RCA = right coronary artery; RFR = relative flow reserve; sMBF = stress myocardial blood flow.

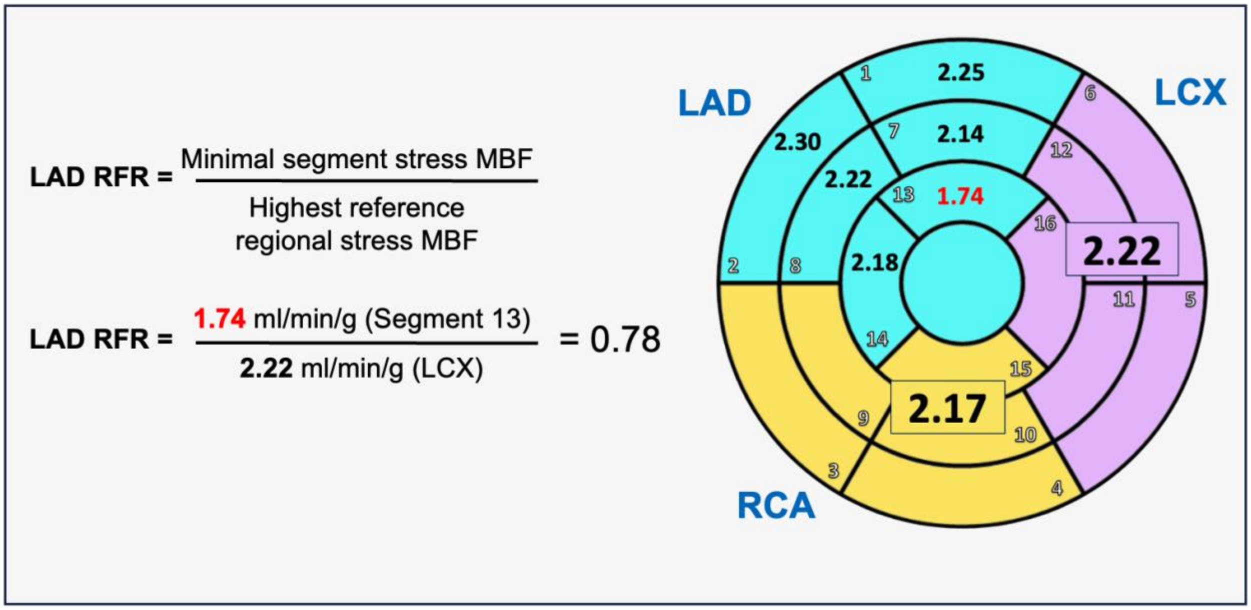

**Supplementary Figure 2. Unadjusted per-vessel ROC Analysis comparing RFR to other PET metrics in the diagnosis of obstructive CAD in all vessels.** The analysis was performed on 693 vessels from 231 patients. The area under the curve (AUC) and 95% confidence intervals were derived from clustered bootstrapping to account for within-patient correlation. All PET parameters represent regional (per-vessel) values. MFR = myocardial flow reserve; RFR = relative flow reserve; sTPD = stress total perfusion deficit.

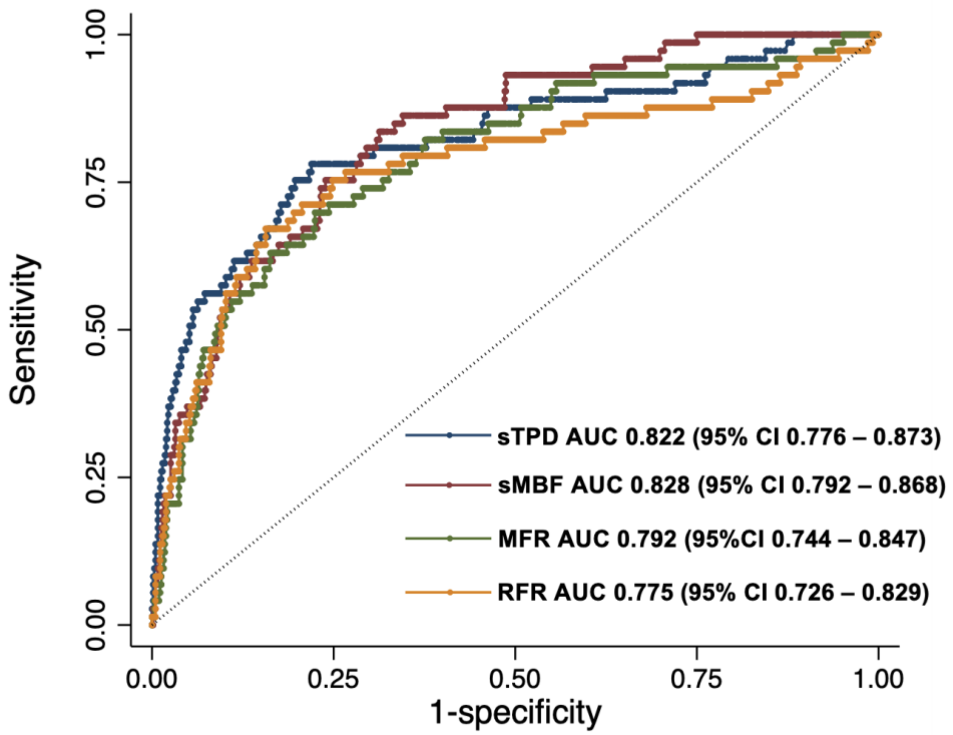

**Supplementary Figure 3. Interaction of per-vessel RFR with (A) stress MBF and (B) stress TPD in predicting the probability of obstructive CAD.** Predicted probabilities were derived from a generalized estimating equations model based on 693 vessels from 231 patients. The model, which accounted for within-patient correlation, was adjusted for age, sex, and BMI and treated RFR as a continuous variable. For visualization purposes, RFR is stratified into the four discrete groups shown. MBF = myocardial blood flow; RFR = relative flow reserve; TPD = total perfusion deficit.

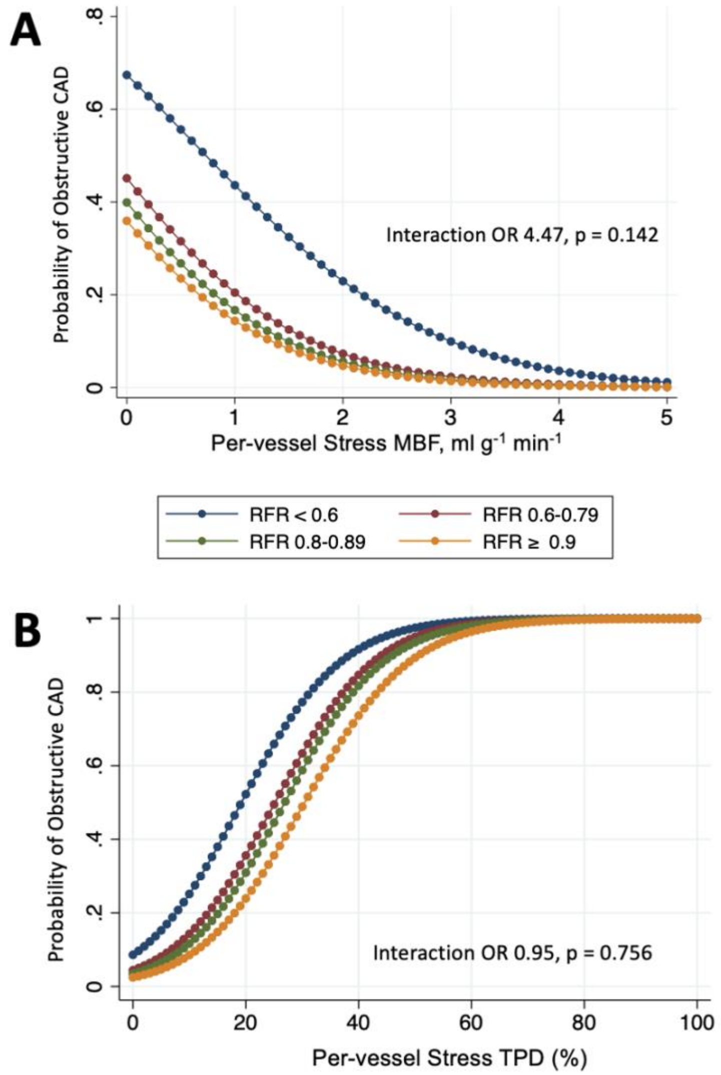

**Supplementary Figure 4. Discrimination of Obstructive and Nonobstructive CAD in Vessels with Reduced Stress MBF using MFR and RFR.** Per-vessel adjusted ROC analysis comparing MFR alone versus stress MFR and RFR for the diagnosis of obstructive CAD. The analysis, based on 346 vessels from 143 patients, included adjustment for age, BMI, and sex, with the p-value and 95% confidence intervals derived from clustered bootstrapping. MBF = myocardial blood flow; MFR = myocardial flow reserve; RFR = relative flow reserve.

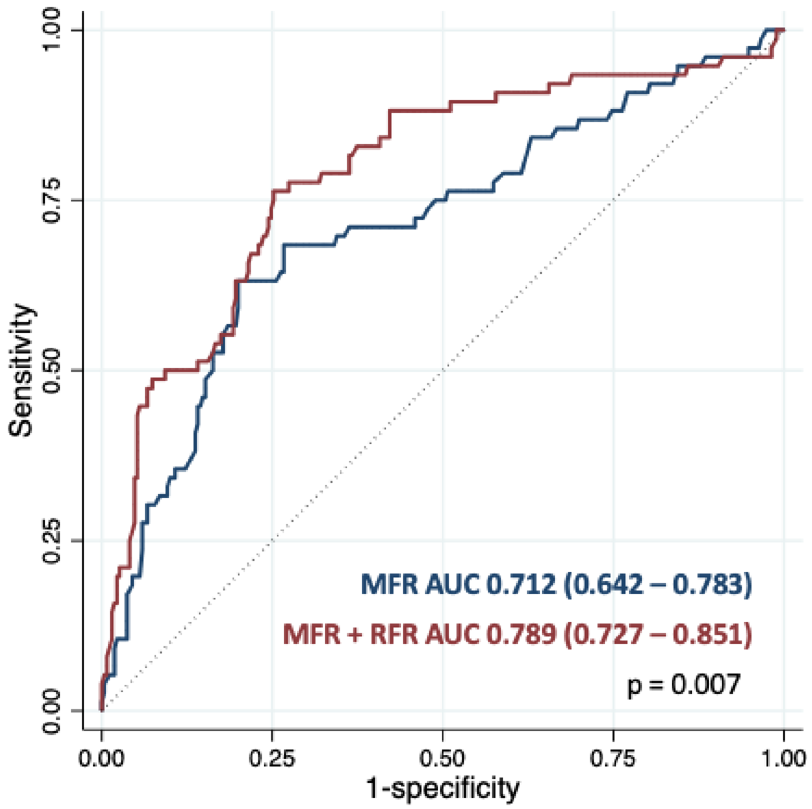

**Supplementary Table 1. Per-patient PET and ICA Results.**

| | ALL (n = 231) | Nonobstructive CAD (n = 620) | Obstructive CAD (n = 73) | p-value |
|---|---|---|---|---|
| **Global sMBF (ml/min/g)** | 2.19 (1.71, 2.77) | 2.39 (1.87, 2.92) | 1.69 (1.32, 2.02) | <0.0001 |
| **LAD sMBF (ml/min/g)** | 2.20 (1.64, 2.69) | 2.23 (1.79, 2.75) | 1.47 (0.89, 1.93) | <0.0001 |
| **LCX sMBF (ml/min/g)** | 2.45 (1.87, 3.03) | 2.51 (1.99, 3.11) | 1.59 (1.39, 2.01) | <0.0001 |
| **RCA sMBF (ml/min/g)** | 2.01 (1.55, 2.64) | 2.21 (1.69, 2.77) | 1.21 (1.06, 1.40) | <0.0001 |
| **Global MFR** | 2.92 (2.28, 3.67) | 3.15 (2.46, 3.94) | 2.37 (1.89, 2.98) | <0.0001 |
| **LAD MFR** | 2.93 (2.28, 3.75) | 2.98 (2.35, 3.78) | 2.23 (1.59, 3.01) | 0.002 |
| **LCX MFR** | 2.95 (2.23, 3.65) | 3.01 (2.36, 3.72) | 2.04 (1.51, 2.65) | <0.0001 |
| **RCA MFR** | 2.83 (2.15, 3.79) | 3.00 (2.34, 3.90) | 1.76 (1.56, 2.11) | <0.0001 |
| **Global sTPD %** | 9.3 (4.1, 18.0) | 7.2 (3.3, 13.4) | 21.6 (12.7, 29.2) | <0.0001 |
| **LAD sTPD %** | 2.4 (1.1, 6.9) | 2.2 (1.1, 5.5) | 15.3 (4.7, 27.5) | <0.0001 |
| **LCX sTPD %** | 1.4 (0.3, 3.4) | 1.3 (0.3, 3.1) | 4.9 (2.0, 13.1) | 0.0001 |
| **RCA sTPD %** | 2.1 (0.7, 5.3) | 1.7 (0.6, 4.0) | 12.4 (7.2, 16.8) | <0.0001 |

Values are median (interquartile range) or number (%). Obstructive CAD refers to ≥1 vessel territory with ≥ 70% stenosis (or LM ≥ 50%). ICA = invasive coronary angiogram; LAD = the left anterior descending artery; LCX = left circumflex artery; MFR = myocardial flow reserve; PET = positron emission tomography; RCA = right coronary artery; sMBF = stress myocardial blood flow; sTPD = stress total perfusion deficit.
